# Clinician-Led Remote Hypertension Monitoring and Blood Pressure Control in a Majority-Minority Primary Care Cohort: Racial Disparities and Equity Implications

**DOI:** 10.64898/2026.07.12.26357888

**Authors:** Ryan Mackey, Ronak Bharucha, Andrew Monte, Andrew Spitznogle, Ajay Baindur, Daim Sardar, Xavier Zonna, Sayuri Gurusinghe, Erik Beeler, Ali Khan, Yilin Xu, Rebekah J. Walker, Ellen Rich

## Abstract

**Background:** Racial and ethnic minority populations face disproportionate rates of uncontrolled blood pressure (BP) and hypertension-related mortality. Remote hypertension monitoring (RHM) with active clinician-led medication titration has shown promise for improving BP control, but real-world evidence in majority-minority primary care settings remains limited.

**Methods:** This retrospective cohort study (January 2022–December 2024) enrolled adults with hypertension in a Bluetooth-integrated RHM program at a single urban academic primary care clinic. Of 550 patients enrolled, 503 with evaluable follow-up data were included. Patients transmitted daily home BP readings; clinicians reviewed readings monthly and titrated anti-hypertensive regimens per 2017 ACC/AHA guidelines. BP control was assessed at baseline and 3, 6, and 9 months. Factors associated with longitudinal BP control were examined using multivariable generalized estimating equations (GEE), with outcomes defined as strict control (<130/80 mmHg), at-least-moderate control (<140/90 mmHg), and uncontrolled (>140/90 mmHg).

**Results:** Among 503 participants (mean age 58.3 [SD 12.1] years; 63.6% African American; 52.9% male), BP control increased from 10.1% at baseline to 37.1% at 9 months. Each additional month of enrollment was associated with reduced odds of uncontrolled BP (adjusted odds ratio [aOR] 0.82; 95% CI, 0.80–0.85; P<.001). White race was associated with lower odds of uncontrolled BP versus African American race (aOR 0.57, at-least-moderate control; aOR 0.40, strict control; both P<.001). Male sex (aOR 1.46; P=.02) and congestive heart failure (aOR 2.09, strict control; aOR 2.05, at-least-moderate control; both P<.05) were associated with higher odds of uncontrolled BP.

**Conclusion:** Bluetooth-integrated RHM with active clinician-led medication titration was associated with a nearly 4-fold increase in BP control over 9 months in a majority-minority primary care population. Persistent within-program racial disparities underscore the need for equity-centered strategies beyond technology adoption alone. Prospective studies with concurrent usual-care comparators are needed to establish causal inference.

**What is Known:**

- Remote hypertension monitoring combined with active pharmacist- or clinician-led medication titration reduces blood pressure more effectively than passive monitoring or usual care alone, as demonstrated in randomized trials conducted predominantly in White, non-minority populations.
- Black Americans bear a disproportionate burden of hypertension, with earlier onset, greater severity, and higher rates of hypertensive end-organ damage compared with other racial and ethnic groups, and persistent disparities in BP control remain despite similar awareness and treatment rates.

**What the Study Adds:**

- In one of the largest single-center cohort studies of physician-led remote hypertension monitoring in a majority-African American urban primary care population, BP control rates increased nearly 4-fold (10.1% to 37.1%) over 9 months, with each month of enrollment independently associated with an 18% reduction in the odds of uncontrolled BP.
- Despite overall program efficacy, significant within-program racial disparities persisted — White patients had 43%–60% lower odds of uncontrolled BP compared with African American patients — and male sex and congestive heart failure were independently associated with failure to achieve BP control targets, identifying high-priority subgroups for augmented intervention strategies.

## Introduction

Hypertension is the leading modifiable risk factor for cardiovascular disease and stroke, affecting approximately 116 million U.S. adults.^1,2^ Despite the availability of effective pharmacotherapy, achieving goal blood pressure in traditional primary care settings remains challenging, largely due to clinical inertia and infrequent monitoring.^3^ The problem is particularly acute in racial and ethnic minority populations. Black Americans have the highest prevalence of hypertension among all U.S. racial/ethnic groups and are 40% more likely to have uncontrolled blood pressure compared with non-Hispanic White Americans despite comparable rates of hypertension awareness and treatment.^4^ They are five times more likely to die from hypertension-related causes and experience disproportionate rates of hypertensive end-organ damage.^4,5^

Remote hypertension monitoring (RHM) has emerged as an evidence-based strategy to overcome clinical inertia by enabling continuous BP surveillance and structured medication titration outside the office setting.^7^ A landmark randomized trial by Margolis et al. demonstrated significant BP reduction with home telemonitoring combined with pharmacist management.^7^ More recently, however, a 2024 randomized clinical trial by Mehta et al. found that remote BP monitoring alone without active medication titration did not significantly improve control compared with usual care, highlighting the importance of the clinician-directed intervention component in producing meaningful BP reductions.^8^

Despite growing evidence for RHM efficacy, few large-scale implementations have been evaluated specifically in underserved, majority-minority primary care settings, which face compounding barriers including lower health literacy, limited digital access, and structural social determinants of health.^4,9^ This study evaluates the association between a Bluetooth-integrated remote hypertension monitoring program and longitudinal BP control and identifies clinical and demographic factors associated with control in a cohort that is majority African American. We hypothesized that program enrollment would be associated with significantly improved BP control and that certain demographic and comorbidity subgroups would exhibit differential responses warranting targeted intervention. The program utilized the Brook Remote Care platform (Brook.ai), which integrates cellular-connected BP cuffs with AI-assisted clinical dashboards to support continuous between-visit monitoring and facilitate clinician-directed medication titration.

This study is informed by the National Institute on Minority Health and Health Disparities (NIMHD) Research Framework, which situates race and ethnicity as social rather than biological constructs and directs attention to the interplay of behavioral, physical/built environment, sociocultural, and healthcare system factors—operating at individual, interpersonal, community, and societal levels—that produce and sustain health disparities. Consistent with this framework, race/ethnicity is analyzed here as a marker of structural and social exposure rather than a proxy for biological difference, and study factors such as healthcare access, comorbidity burden, and program engagement are interpreted within this broader context of structural determinants.^6^

## Methods

### Study Design and Setting

This retrospective single-center cohort study was conducted at an urban academic internal medicine clinic between January 2022 and December 2024. The study was approved by the University at Buffalo Institutional Review Board (IRB ID: STUDY00010652). Reporting adheres to the Strengthening the Reporting of Observational Studies in Epidemiology (STROBE) guidelines.^10^

### Inclusion and Exclusion Criteria

#### Inclusion criteria

1. Adult patients aged ≥18 years; (2) established diagnosis of hypertension with at least one prior clinic visit; (3) enrollment in the remote BP monitoring program during the study period; (4) Evaluable remote BP data at one or more follow-up intervals.

#### Exclusion criteria

Age <18 years; (2) inability to operate the BP cuff independently due to physical or cognitive impairment; (3) discharge from the remote hypertension program due to lack of transmitted blood pressure readings or failure to attend follow up visits.

### Intervention

Enrolled patients were provided a validated, Bluetooth-capable automated BP cuff and instructed to perform daily home BP measurements. Readings were automatically transmitted to a secure, HIPAA-compliant database accessible to the clinical team. Registered nurses managed two components of the care model. First, nurses conducted structured onboarding visits with each newly enrolled patient to provide education on device use, BP self-monitoring technique, and program expectations. Second, nurses monitored incoming BP transmissions on the remote monitoring dashboard and served as the initial point of contact when transmitted readings met pre-specified alert thresholds. When a threshold was triggered, nurses contacted patients directly to assess for symptoms, medication adherence, and contributing factors; findings from these interactions were then communicated to the treating physician. Nurses also responded to patient-initiated contact between scheduled virtual visits for BP-related concerns. Nursing involvement was integrated into existing ambulatory care workflows. Monthly BP summaries were generated and reviewed with patients during structured virtual encounters (telehealth video or telephone) conducted by resident or attending physicians. At each virtual visit, clinicians reviewed BP trends, performed medication titration per a hypertension management protocol aligned with 2017 ACC/AHA guidelines,^1^ and provided lifestyle modification counseling (dietary sodium reduction, exercise, smoking cessation, alcohol moderation, etc.). Medication changes were documented in the electronic medical record.

### Data Collection

Deidentified demographic data, medical history, laboratory values, and medication regimens were extracted from the electronic medical record (EMR) for all enrolled patients. Extracted demographic variables included age, sex, and self-reported race/ethnicity (categorized as African American, White, or Other). Clinical and comorbidity variables included baseline systolic and diastolic blood pressure; number of concurrent anti-hypertensive medications (0, 1, 2, or ≥3); presence of type 2 diabetes mellitus; congestive heart failure and left ventricular ejection fraction category (normal [>49%], reduced/moderately reduced [<50%], or missing); chronic kidney disease; hyperlipidemia; coronary artery disease; atrial fibrillation; prior transient ischemic attack, stroke, or subarachnoid hemorrhage; obesity (body mass index >30 kg/m²); and smoking status (current, former, or never). Laboratory values extracted included total cholesterol, high-density lipoprotein (HDL) cholesterol, low-density lipoprotein (LDL) cholesterol, and triglycerides. Medication use variables included statin therapy, anticoagulation therapy, and aspirin therapy. A complete summary of these baseline variables, stratified by blood pressure control status, is provided in the supplemental, Table S1.

### Outcome Assessment

Baseline BP values were taken directly from the electronic medical record at the time of program enrollment. Blood pressure control status at each follow-up time point (3, 6, and 9 months) was determined using the monthly summary reports generated by the Brook Remote Care (Brook.ai) platform, which aggregated daily home readings transmitted from each patient’s cellular-connected Bluetooth cuff. These reports served as the source data for outcome ascertainment at each follow-up time point. Average BP readings and anti-hypertensive regimens were recorded at enrollment (baseline), 3, 6, and 9 months. BP was categorized as: controlled (<130/80 mmHg), moderately controlled (130–139/80–89 mmHg), or uncontrolled (≥140/90 mmHg).

### Statistical Analysis

Generalized Estimating Equations (GEE) with an exchangeable working correlation structure were used to model the effect of time on the probability of uncontrolled BP, accounting for the within-individual correlation of repeated measures. Models were estimated using R version 4.4.1 and statistical significance was defined as a two-sided P<.05. The fully adjusted model includes both time-invariant (e.g., age, sex, race) and time-variant (e.g., medication use, comorbidities, smoking status, and time) variables to examine their associations with the probability of a dichotomized blood pressure outcome using GEE. Two dichotomized BP outcome models were implemented. Model 1 examined controlled vs moderately controlled and uncontrolled subsets (strict control), and model 2 examined controlled and moderately controlled vs uncontrolled subsets (at-least-moderate control). Adjusted odds ratios (aOR) and 95% confidence intervals (CI) are reported. Given the 27% attrition rate over 9 months, a complete-case sensitivity analysis was performed and results were consistent with the primary model. No imputation was performed for missing follow-up readings.

## Results

### Cohort Characteristics

Of 550 patients enrolled, 503 had evaluable data at one or more follow-up intervals and were included in the primary analysis. The remaining 47 patients had no BP transmissions after enrollment and were excluded. The mean age was 58.3 years (SD 12.1). The cohort was 63.6% African American, 52.9% male, 28.8% White, and 7.6% other race/ethnicity. Comorbidities included, but are not limited to, type 2 diabetes mellitus (31.8%), congestive heart failure (7.6%), chronic kidney disease (34.6%), obesity (59.0%), and current smoking (23.9%). A complete list of baseline cohort characteristics can be found in Table S1. At baseline, only 10.1% of patients (n=51) had controlled BP (<130/80 mmHg). Select baseline characteristics are summarized in Table 1.

**Table 1.**
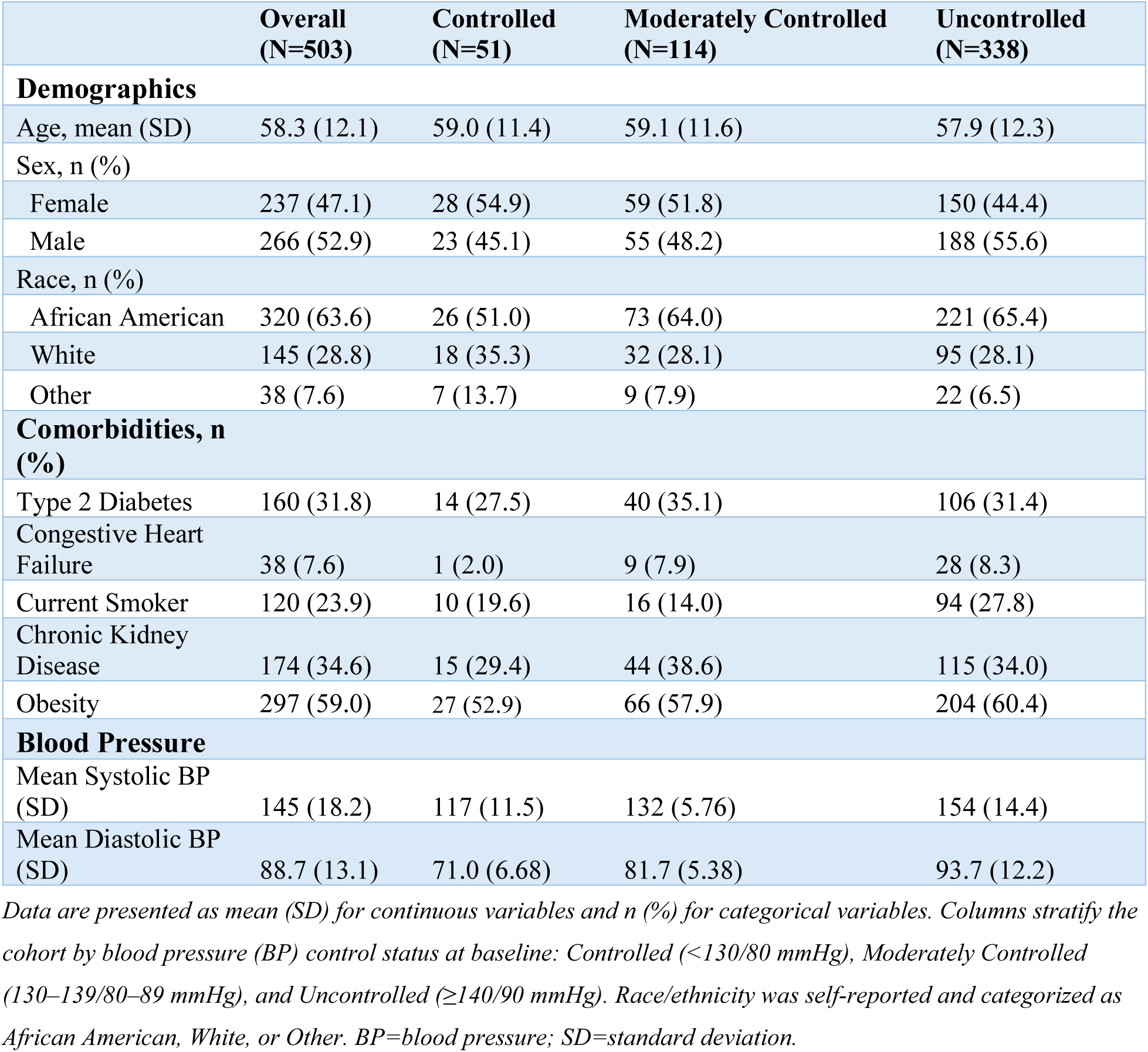
Baseline Characteristics of the Study Cohort (N=503)

### Efficacy of Remote Monitoring

Blood pressure control improved significantly over the study period. By month 9, 37.1% of patients had controlled BP, compared with 10.1% at baseline — a nearly four-fold increase. Longitudinal GEE analysis demonstrated that each additional month of program enrollment was associated with an 18% reduction in the odds of uncontrolled BP (fully adjusted OR 0.82; 95% CI, 0.80–0.85; P<.001). As illustrated in Figure 1, the proportion achieving BP control increased steadily throughout the study period, while the proportion of uncontrolled patients declined by more than half. Detailed summary data for each follow-up visit are provided in Tables S2a, S2b, and S2c (3, 6, and 9 months, respectively).

**Figure 1:**
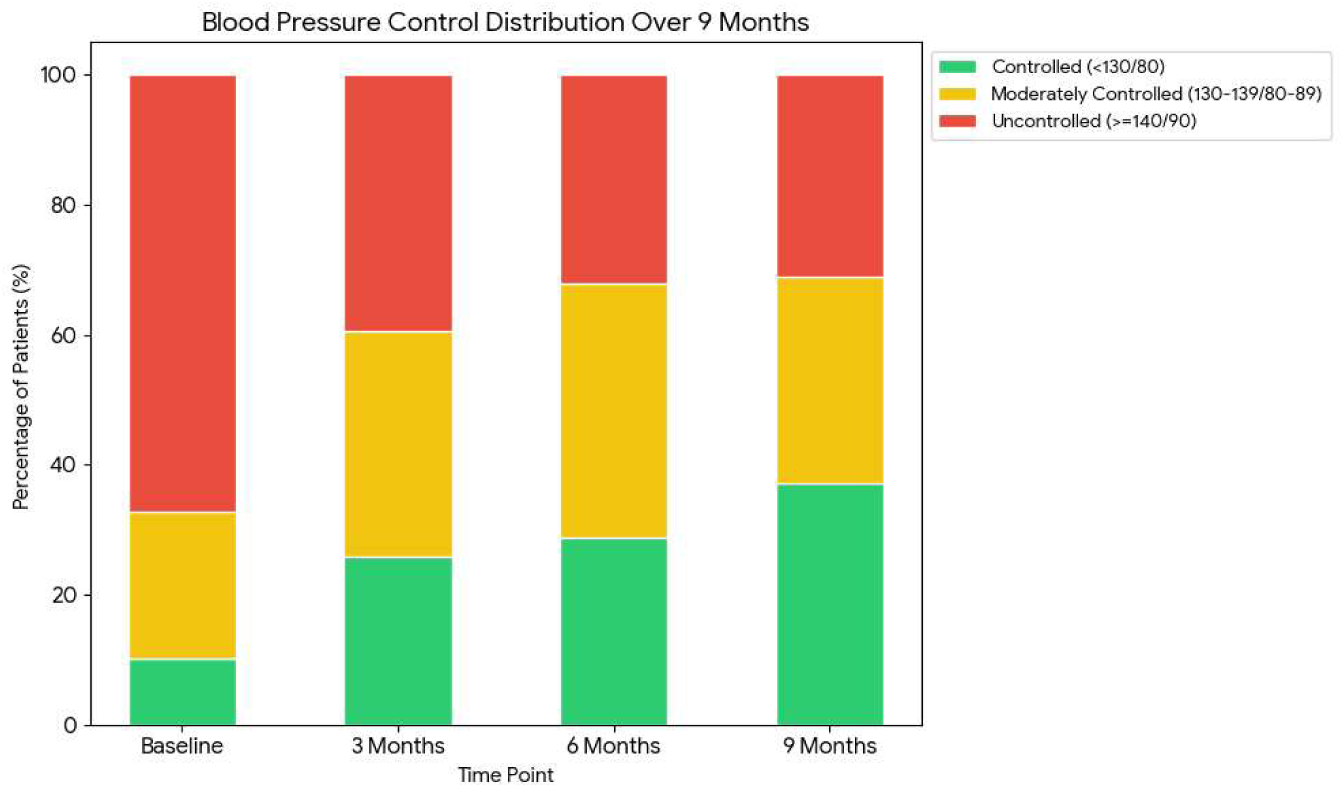
Longitudinal Distribution of Blood Pressure Control over 9 Months. Stacked bar chart representing the categorical distribution of blood pressure control for the cohort from enrollment to 9 months. The proportion of patients achieving BP control increased from 10.1% at baseline to 37.1% at the 9-month follow-up.

### Factors Associated with BP Control

Longitudinal analysis using Generalized Estimating Equations (GEE) identified several significant clinical and demographic factors associated with blood pressure control. These associations varied depending on whether the outcome was defined as achieving “strict control” (<130/80 mmHg), as generated by the model 1 (Table S3), or achieving “at-least-moderate control” (<140/90 mmHg), as generated by model 2 (Table S4).

#### Factors Associated with Strict Control (<130/80 mmHg)

In the fully adjusted model comparing patients with controlled BP against those who were moderately controlled or uncontrolled, time in program was significantly associated with BP control. Each month of enrollment was associated with a 16% reduction in the odds of being uncontrolled/moderately controlled (aOR 0.84; 95% CI, 0.81–0.87; P<.001). Several patient-level demographic characteristics were significantly associated with BP control. Older age was associated with modestly lower odds of remaining uncontrolled or moderately controlled (aOR 0.98 per year; 95% CI, 0.97–1.00; P=.01). White patients had significantly higher odds of achieving strict control compared to African American patients (aOR 0.40; 95% CI, 0.28–0.57; P<.001). Male sex was associated with 46% higher odds of remaining in the uncontrolled or moderately controlled group (aOR 1.46; 95% CI, 1.06–2.02; P=.02). Certain comorbidities were significantly associated with BP control. Patients with type 2 diabetes were more likely to achieve strict control compared to those without (aOR 0.65; 95% CI, 0.46–0.93; P=.02). Conversely, congestive heart failure was associated with nearly twice the odds of failing to reach strict control (aOR 2.09; 95% CI, 1.15–3.79; P=.02).

#### Factors Associated with At Least Moderate Control (<140/90 mmHg)

When analyzing the outcome of controlled or moderately controlled vs. uncontrolled BP, time in the program was significantly associated with BP control. Monthly enrollment remained highly significant, with an 18% reduction in the odds of being uncontrolled (aOR 0.82; 95% CI, 0.80–0.85; P<.001). Older age was similarly associated with modestly lower odds of being uncontrolled (aOR 0.98 per year; 95% CI, 0.97–0.99; P<.01). The disparity tied to race remained significant, with White patients having lower odds of being uncontrolled compared to African American patients (aOR 0.57; 95% CI, 0.41–0.78; P<.001). Certain comorbidities were significantly associated with BP control. Current smoking was associated with higher odds of uncontrolled BP at the borderline of conventional significance (aOR 1.45; 95% CI, 1.01–2.10; P=.05) and should be interpreted with caution. Congestive heart failure was associated with nearly twice the odds of failing to reach at-least-moderate control (aOR 2.05; 95% CI, 1.05–4.02; P=.04).

## Discussion

This retrospective cohort study demonstrates that a Bluetooth-integrated remote hypertension monitoring program with active clinician-led medication titration was associated with a nearly four-fold increase in BP control rates — from 10.1% to 37.1% — over 9 months in a real-world, majority-minority primary care population. To our knowledge, this represents one of the larger single-center evaluations of RHM efficacy specifically in an urban, majority-African American cohort.

The magnitude of improvement observed in this study compares favorably with prior RHM trials. The landmark 2013 JAMA randomized controlled trial by Margolis et al. demonstrated significant systolic BP reductions with home telemonitoring combined with pharmacist management in a predominantly White primary care population; our findings extend this evidence to a majority-minority cohort with physician-led titration.^7^ A 2021 meta-analysis of 32 high-quality remote BP monitoring studies found a pooled standardized mean difference of 0.51 (95% CI, 0.34–0.68) in systolic BP reduction compared with usual care, supporting the broad efficacy of RHM.^11^ Notably, a 2024 randomized clinical trial published in *JAMA Network Open* by Mehta et al. found that remote monitoring alone — without active medication titration — did not significantly improve BP control compared with usual care.^8^ This null result directly underscores the importance of the structured virtual visit and medication titration component embedded in our program, rather than passive data transmission alone, as the driver of BP improvement.

### Dichotomized Models: Strict and At Least Moderate Control

The comparison of the two dichotomized models reveals critical nuances in patient management within a remote monitoring framework. The variable of time shows a slightly stronger effect in the moderate control model (18% vs 16% reduction), suggesting the program is highly effective at moving the most “at-risk” (uncontrolled) patients into a safer range (moderately controlled), even if they do not yet meet the strict AHA goal of < 130/80.

#### Racial Disparities in Blood Pressure Control

The racial disparity in BP control within this program merits careful consideration given the study’s stated focus on health equity. Depending on the BP control threshold applied, White patients had 43% lower odds of remaining uncontrolled relative to at-least-moderate control (aOR 0.57, Model 2) and 60% lower odds of failing to achieve strict control (aOR 0.40, Model 1) compared with African American patients. These findings are consistent with well-established national patterns. Black Americans develop hypertension at an earlier age, with greater severity, and experience faster disease progression than their White counterparts. Notably, the “Other” race/ethnicity category (7.6% of the cohort) is not further characterized in this dataset, limiting interpretation for Hispanic/Latino, Asian, and additional minority subgroups who may face distinct barriers within remote monitoring programs.^4^ Structural racism, experiences of discrimination in health care, and socioeconomic and environmental inequities have been documented as contributors to persistent BP disparities beyond pharmacologic treatment alone.^4,5^

Equity-centered interventions that pair remote monitoring with community health workers, and standardized medication titration protocols have demonstrated promise in closing racial gaps in BP control among Black Americans.^4^ The HHS Office of Minority Health and the American Heart Association’s National Hypertension Control Initiative has specifically targeted integration of remote BP monitoring technology into HRSA-funded minority-serving health centers as a strategy to reduce these disparities.^12^ Future iterations of this program should consider layering culturally conscious patient education and social determinants of health screening alongside BP monitoring, as remote technology alone appears insufficient to fully eliminate racial disparities in BP control.

#### Association of Male Sex with Poor BP Control

Male sex was associated with significantly higher odds of failing to achieve strict BP control (<130/80 mmHg) in this cohort (aOR 1.46; P=.02, Model 1). Notably, this association did not reach statistical significance for at-least-moderate control (aOR 1.19; P=.24, Model 2), suggesting that male sex was associated with lower odds of achieving the most stringent BP targets but not associated with a lack of improvement in BP. This finding is consistent with literature demonstrating that women with hypertension generally have higher rates of BP control and medication adherence than men.^13^ A review of Swedish primary care registry data found that BP control was better in women than men throughout most of the lifespan.^13^ In the setting of resistant hypertension specifically, male sex has been identified as an independent risk factor for end-organ damage, including higher prevalence of left ventricular hypertrophy and proteinuria, and for adverse cardiovascular outcomes including myocardial infarction and all-cause mortality.^14^

Mechanistic contributors to this difference likely include differences in the renin-angiotensin-aldosterone system (RAAS) as testosterone promotes higher RAAS activity and vasomotor tone in men, potentially increasing resistance to standard anti-hypertensive regimens.^15^ Behavioral factors including lower engagement with preventive care visits, lower medication adherence, and lower responsiveness to remote monitoring modalities among men are also well-documented.^13^ Future program iterations should explore stratified outreach strategies, including more frequent virtual touchpoints and targeted health literacy messaging for male patients, to alleviate this disparity.

#### Congestive Heart Failure and Blood Pressure Control

Congestive heart failure emerged as the factor most strongly associated with poor BP control across both models — more than doubling the odds of failing to achieve strict control (aOR 2.09; 95% CI, 1.15–3.79; Model 1) and at-least-moderate control (aOR 2.05; 95% CI, 1.05–4.02; Model 2). This finding reflects the unique complexity of managing hypertension in the setting of heart failure. Volume fluctuations inherent to CHF and the associated adjustments in diuretic therapy can produce dynamic BP variability that does not reflect stable, treatment-refractory hypertension.^16^ Residual pulmonary congestion persists in 30–40% of patients after acute decompensation,^17^ and diuretic resistance, present in up to 30% of patients with decompensated heart failure, further complicates hemodynamic management.^18^

#### Type 2 Diabetes and Better Blood Pressure Control

Patients with type 2 diabetes mellitus were significantly more likely to achieve a strict BP control of <130/80 (aOR 0.65). While we attribute this finding in part to higher baseline health system engagement — more frequent laboratory checks, specialist co-management, and insurance-driven performance metrics — several pharmacologic factors likely contribute. SGLT2 inhibitors and GLP-1 receptor agonists, increasingly used as first-line agents in type 2 diabetes, carry meaningful anti-hypertensive effects independent of their glycemic actions.^19^ This potential pharmacologic confounding should be acknowledged when interpreting the diabetes–BP control association, and future studies should capture anti-hypertensive medication class as a covariate in GEE models.

#### Antihypertensive Medication Count as a Covariate

The number of anti-hypertensive medications was included as a covariate in the adjusted GEE models; however, medication count is itself a downstream product of the intervention (active medication titration) and therefore partially endogenous. The counterintuitive finding that 2 or more anti-hypertensive agents were associated with higher odds of remaining uncontrolled in Model 2 most likely reflects confounding by indication — patients requiring more medications are those with more resistant hypertension — rather than a causal effect of medication count on BP outcomes. Future iterations of these models should consider moving medication count to a sensitivity analysis or mediation analysis framework.

### Clinical Implications

Taken together, these findings support the scalability of RHM as a tool for improving hypertension management in real-world, high-risk primary care settings. The program’s active clinician-led titration component, rather than passive monitoring alone, appears to be the critical driver of improvement, consistent with the 2024 Mehta et al. null result for monitoring-only models.^8^ The infrastructure supporting this program, including the Brook Health remote monitoring platform and nurse-led patient outreach, represents one replicable implementation model for institutions seeking to operationalize structured RHM in similar majority-minority primary care settings. National initiatives such as the HHS/AHA National Hypertension Control Initiative have recognized this and are funding implementation of structured RHM specifically in minority-serving federally qualified health centers.^12^ However, our program’s racial disparities suggest that technology adoption alone does not eliminate systemic inequities, and that equity-centered multidisciplinary interventions remain necessary.

### Limitations

This study has several important limitations. First, the 27% attrition rate over 9 months introduces the risk of attrition bias if patients who discontinued participation had systematically different BP trajectories than completers, although GEE models are robust to informative missingness under the missing-at-random assumption. Patients who dropped out were not formally compared to completers, and a sensitivity analysis using complete-case methods yielded results consistent with the primary model. Second, as a single-center retrospective cohort study without a concurrent usual-care control group, causal attribution of BP improvement to the RHM program cannot be established with certainty. With 67.2% of patients uncontrolled at baseline (mean SBP 154 mmHg in that group), regression to the mean alone would be expected to produce some degree of improvement independent of any intervention. Secular trends in anti-hypertensive prescribing during the 2022–2024 study period and increased healthcare engagement attributable to program enrollment rather than remote monitoring technology specifically cannot be excluded as contributing factors. Third, selection bias is possible: patients who enrolled in remote monitoring and were provided Bluetooth-capable devices may be systematically more engaged than those who declined or lacked digital access, limiting generalizability to populations with lower digital literacy. Fourth, the study did not collect data on socioeconomic status, educational attainment, neighborhood-level social determinants of health, or insurance type — important confounders in a study with an explicit health equity focus. Fifth, medication adherence was not measured; the study quantifies medication titration decisions but cannot determine whether patients took prescribed medications as directed, a critical confounder of BP outcomes. Sixth, the “Other” race/ethnicity category (7.6%) is not further specified in the dataset, limiting interpretation of results for Hispanic/Latino, Asian, and other minority subgroups. Future studies should prospectively collect self-reported race and ethnicity using disaggregated categories (e.g., Hispanic/Latino, Asian, and other specific subgroups) rather than a composite “Other” category, to enable more precise characterization of disparities within remote hypertension monitoring programs. Finally, the single-center design at an urban academic institution limits generalizability to community health centers and rural settings with different resources and patient populations. Additionally, the program was implemented using a commercial platform (Brook.ai); whether comparable outcomes would be achieved using alternative remote monitoring systems or internally developed infrastructure remains unknown and limits direct generalizability of the implementation model.

## Conclusion

A Bluetooth-integrated remote hypertension monitoring program with active clinician-led medication titration was associated with a nearly four-fold increase in BP control rates over 9 months in a majority-minority urban primary care cohort. Given the retrospective single-center design without a concurrent usual-care control group, causal attribution of this improvement to the RHM program cannot be established with certainty. Persistent within-program racial disparities in BP control underscore the need for equity-centered supplementary strategies beyond technology adoption alone. Targeted interventions for male patients and those with congestive heart failure — including more intensive virtual follow-up and integration with disease-specific management programs — may further improve outcomes. Current smoking was associated with failure to achieve at-least-moderate BP control at a borderline significance level and warrants further investigation as an intervention target. Future research should evaluate cost-effectiveness, long-term impact on major adverse cardiovascular events, and the additive benefit of culturally congruent patient engagement strategies in this population.

## Acknowledgments

None

## Data Availability

The data that support the findings of this study are not publicly available due to institutional and patient privacy restrictions but may be made available from the corresponding author upon reasonable request, pending institutional approval and execution of a data use agreement.

## Abbreviations List

ACC/AHA: American College of Cardiology/American Heart Association
aOR: Adjusted odds ratio
ANOVA: Analysis of variance
BMI: Body mass index
BP: Blood pressure
CHF: Congestive heart failure
CI: Confidence interval
DBP: Diastolic blood pressure
EMR: Electronic medical record
GEE: Generalized estimating equations
GLP-1: Glucagon-like peptide-1
HDL: High-density lipoprotein
HHS: U.S. Department of Health and Human Services
IRB: Institutional Review Board
LDL: Low-density lipoprotein
NIMHD: National Institute on Minority Health and Health Disparities
RAAS: Renin-angiotensin-aldosterone system
RHM: Remote hypertension monitoring
SBP: Systolic blood pressure
SD: Standard deviation
SGLT2: Sodium-glucose cotransporter-2
STROBE: Strengthening the Reporting of Observational Studies in Epidemiology

## Supplemental Material

**Table S1:** Summary of Baseline Data.

|  | **Controlled** | **Moderately Controlled** | **Uncontrolled** | **Overall** | **p-value <sup>1</sup>** |
| --- | --- | --- | --- | --- | --- |
|  | **(N=51)** | **(N=114)** | **(N=338)** | **(N=503)** |  |
| **Age** |  |  |  |  | 0.6 |
| Mean (SD) | 59.0 (11.4) | 59.1 (11.6) | 57.9 (12.3) | 58.3 (12.1) |  |
| Median [Min, Max] | 61.0 [33.0, 79.0] | 60.0 [27.0, 86.0] | 58.0 [28.0, 87.0] | 59.0 [27.0, 87.0] |  |
| **Sex** |  |  |  |  | 0.2 |
| Female | 28 (54.9%) | 59 (51.8%) | 150 (44.4%) | 237 (47.1%) |  |
| Male | 23 (45.1%) | 55 (48.2%) | 188 (55.6%) | 266 (52.9%) |  |
| **Race** |  |  |  |  | 0.25 |
| African-American | 26 (51.0%) | 73 (64.0%) | 221 (65.4%) | 320 (63.6%) |  |
| Other | 7 (13.7%) | 9 (7.9%) | 22 (6.5%) | 38 (7.6%) |  |
| White | 18 (35.3%) | 32 (28.1%) | 95 (28.1%) | 145 (28.8%) |  |
| **SBP** |  |  |  |  | <0.001 |
| Mean (SD) | 117 (11.5) | 132 (5.76) | 154 (14.4) | 145 (18.2) |  |
| Median [Min, Max] | 122 [78.0, 129] | 133 [113, 139] | 150 [123, 215] | 144 [78.0, 215] |  |
| **DBP** |  |  |  |  | <0.001 |
| Mean (SD) | 71.0 (6.68) | 81.7 (5.38) | 93.7 (12.2) | 88.7 (13.1) |  |
| Median [Min, Max] | 72.0 [50.0, 79.0] | 83.0 [63.0, 89.0] | 93.0 [50.0, 146] | 88.0 [50.0, 146] |  |
| **Number of Anti-Hypertensive Medications** |  |  |  |  | 0.23 |
| 0 | 3 (5.9%) | 15 (13.2%) | 39 (11.5%) | 57 (11.3%) |  |
| 1 | 26 (51.0%) | 36 (31.6%) | 112 (33.1%) | 174 (34.6%) |  |
| 2 | 15 (29.4%) | 36 (31.6%) | 109 (32.2%) | 160 (31.8%) |  |
| 3 or more than 3 | 7 (13.7%) | 27 (23.7%) | 78 (23.1%) | 112 (22.3%) |  |
| **Type 2 Diabetes Mellitus** |  |  |  |  | 0.59 |
| No | 37 (72.5%) | 74 (64.9%) | 232 (68.6%) | 343 (68.2%) |  |
| Yes | 14 (27.5%) | 40 (35.1%) | 106 (31.4%) | 160 (31.8%) |  |
| **Congestive Heart Failure** |  |  |  |  | 0.28 |
| No | 50 (98.0%) | 105 (92.1%) | 310 (91.7%) | 465 (92.4%) |  |
| Yes | 1 (2.0%) | 9 (7.9%) | 28 (8.3%) | 38 (7.6%) |  |
| **Ejection Fraction** |  |  |  |  | 0.35 |
| Normal (>49%) | 42 (82.4%) | 98 (86.0%) | 305 (90.2%) | 445 (88.5%) |  |
| Reduced (<35%)/Moderately Reduced (35%-49%) | 0 (0%) | 4 (3.5%) | 15 (4.4%) | 19 (3.8%) |  |
| Missing | 9 (17.6%) | 12 (10.5%) | 18 (5.3%) | 39 (7.8%) |  |
| **Chronic Kidney Disease** |  |  |  |  | 0.48 |
| No | 36 (70.6%) | 70 (61.4%) | 223 (66.0%) | 329 (65.4%) |  |
| Yes | 15 (29.4%) | 44 (38.6%) | 115 (34.0%) | 174 (34.6%) |  |
| **Hyperlipidemia** |  |  |  |  | 0.46 |
| No | 10 (19.6%) | 21 (18.4%) | 80 (23.7%) | 111 (22.1%) |  |
| Yes | 41 (80.4%) | 93 (81.6%) | 258 (76.3%) | 392 (77.9%) |  |
| **Total Cholesterol** |  |  |  |  | 0.74 |
| Mean (SD) | 182 (51.9) | 176 (41.9) | 178 (43.3) | 178 (43.9) |  |
| Median [Min, Max] | 176 [89.0, 322] | 172 [73.0, 284] | 178 [46.0, 367] | 177 [46.0, 367] |  |
| Missing | 1 (2.0%) | 6 (5.3%) | 24 (7.1%) | 31 (6.2%) |  |
| **HDL** |  |  |  |  | 0.76 |
| Mean (SD) | 55.7 (26.3) | 52.9 (16.4) | 53.2 (24.4) | 53.4 (23.0) |  |
| Median [Min, Max] | 51.0 [23.0, 180] | 49.0 [26.0, 109] | 47.0 [20.0, 274] | 48.0 [20.0, 274] |  |
| Missing | 1 (2.0%) | 6 (5.3%) | 24 (7.1%) | 31 (6.2%) |  |
| **LDL** |  |  |  |  | 0.99 |
| Mean (SD) | 102 (45.4) | 102 (35.2) | 101 (36.7) | 101 (37.3) |  |
| Median [Min, Max] | 95.0 [25.0, 224] | 98.0 [25.0, 212] | 100 [29.0, 250] | 100 [25.0, 250] |  |
| Missing | 1 (2.0%) | 7 (6.1%) | 24 (7.1%) | 32 (6.4%) |  |
| **Triglycerides** |  |  |  |  | 0.13 |
| Mean (SD) | 124 (67.2) | 107 (58.4) | 122 (69.1) | 119 (66.8) |  |
| Median [Min, Max] | 106 [36.0, 407] | 98.5 [20.0, 394] | 102 [8.00, 435] | 102 [8.00, 435] |  |
| Missing | 1 (2.0%) | 8 (7.0%) | 27 (8.0%) | 36 (7.2%) |  |
| **Statin** |  |  |  |  | 0.15 |
| No | 17 (33.3%) | 37 (32.5%) | 141 (41.7%) | 195 (38.8%) |  |
| Yes | 34 (66.7%) | 77 (67.5%) | 197 (58.3%) | 308 (61.2%) |  |
| **Atrial Fibrillation** |  |  |  |  | 0.59 |
| No | 48 (94.1%) | 111 (97.4%) | 324 (95.9%) | 483 (96.0%) |  |
| Yes | 3 (5.9%) | 3 (2.6%) | 14 (4.1%) | 20 (4.0%) |  |
| **Anticoagulation** |  |  |  |  | 0.34 |
| No | 49 (96.1%) | 109 (95.6%) | 312 (92.3%) | 470 (93.4%) |  |
| Yes | 2 (3.9%) | 5 (4.4%) | 26 (7.7%) | 33 (6.6%) |  |
| **Coronary Artery Disease** |  |  |  |  | 0.55 |
| No | 45 (88.2%) | 101 (88.6%) | 287 (84.9%) | 433 (86.1%) |  |
| Yes | 6 (11.8%) | 13 (11.4%) | 51 (15.1%) | 70 (13.9%) |  |
| **Smoker** |  |  |  |  | 0.03 |
| No | 24 (47.1%) | 66 (57.9%) | 154 (45.6%) | 244 (48.5%) |  |
| Yes - Current | 10 (19.6%) | 16 (14.0%) | 94 (27.8%) | 120 (23.9%) |  |
| Yes - Former | 17 (33.3%) | 32 (28.1%) | 90 (26.6%) | 139 (27.6%) |  |
| **Prior Transient Ischemic Attack/Stroke/Subarachnoid Hemorrhage** |  |  |  |  | 0.51 |
| No | 50 (98.0%) | 107 (93.9%) | 319 (94.4%) | 476 (94.6%) |  |
| Yes | 1 (2.0%) | 7 (6.1%) | 19 (5.6%) | 27 (5.4%) |  |
| **Obesity (BMI > 30)** |  |  |  |  | 0.58 |
| No | 24 (47.1%) | 48 (42.1%) | 134 (39.6%) | 206 (41.0%) |  |
| Yes | 27 (52.9%) | 66 (57.9%) | 204 (60.4%) | 297 (59.0%) |  |
| **Aspirin Therapy** |  |  |  |  | 0.88 |
| No | 41 (80.4%) | 88 (77.2%) | 267 (79.0%) | 396 (78.7%) |  |
| Yes | 10 (19.6%) | 26 (22.8%) | 71 (21.0%) | 107 (21.3%) |  |
| <sup>1</sup> The ANOVA test is used for continuous variables, while the Chi-Square test is used for categorical variables. The p-value indicates whether the variable differs across the three blood pressure groups. |  |  |  |  |  |

Data are presented as mean (SD) for continuous variables and n (%) for categorical variables, stratified by blood pressure (BP) control status at baseline (Controlled, Moderately Controlled, Uncontrolled). BMI indicates body mass index; DBP, diastolic blood pressure; HDL, high-density lipoprotein; LDL, low-density lipoprotein; SBP, systolic blood pressure.

**Table S2a:** Summary of 3-Month Visit Data.

|  | Controlled | Moderately Controlled | Uncontrolled | Overall | p-value <sup>1</sup> |
| --- | --- | --- | --- | --- | --- |
|  | (N=121) | (N=162) | (N=184) | (N=467) |  |
| **Age** |  |  |  |  | **0.03** |
| Mean (SD) | 60.2 (11.2) | 58.7 (12.3) | 56.6 (12.2) | 58.3 (12.1) |  |
| Median [Min, Max] | 61.0 [27.0, 83.0] | 60.0 [28.0, 87.0] | 57.0 [29.0, 83.0] | 59.0 [27.0, 87.0] |  |
| **Sex** |  |  |  |  | **0.38** |
| Female | 65 (53.7%) | 78 (48.1%) | 84 (45.7%) | 227 (48.6%) |  |
| Male | 56 (46.3%) | 84 (51.9%) | 100 (54.3%) | 240 (51.4%) |  |
| **Race** |  |  |  |  | **0.03** |
| African-American | 68 (56.2%) | 96 (59.3%) | 129 (70.1%) | 293 (62.7%) |  |
| Other | 6 (5.0%) | 15 (9.3%) | 13 (7.1%) | 34 (7.3%) |  |
| White | 47 (38.8%) | 51 (31.5%) | 42 (22.8%) | 140 (30.0%) |  |
| **SBP** |  |  |  |  | **<0.001** |
| Mean (SD) | 119 (7.54) | 131 (6.30) | 150 (15.4) | 135 (16.9) |  |
| Median [Min, Max] | 121 [88.0, 129] | 132 [104, 139] | 147 [121, 226] | 134 [88.0, 226] |  |
| **DBP** |  |  |  |  | **<0.001** |
| Mean (SD) | 71.0 (6.00) | 81.5 (5.61) | 92.1 (11.2) | 83.0 (11.8) |  |
| Median [Min, Max] | 72.0 [52.0, 79.0] | 82.0 [60.0, 89.0] | 91.0 [66.0, 128] | 82.0 [52.0, 128] |  |
| **Number of Anti-Hypertensive Medications** |  |  |  |  | **0.01** |
| 0 | 12 (9.9%) | 14 (8.6%) | 7 (3.8%) | 33 (7.1%) |  |
| 1 | 33 (27.3%) | 56 (34.6%) | 40 (21.7%) | 129 (27.6%) |  |
| 2 | 41 (33.9%) | 55 (34.0%) | 67 (36.4%) | 163 (34.9%) |  |
| 3 or more than 3 | 35 (28.9%) | 37 (22.8%) | 70 (38.0%) | 142 (30.4%) |  |
| **Type 2 Diabetes Mellitus** |  |  |  |  | **0.3** |
| No | 81 (66.9%) | 118 (72.8%) | 119 (64.7%) | 318 (68.1%) |  |
| Yes | 40 (33.1%) | 44 (27.2%) | 65 (35.3%) | 149 (31.9%) |  |
| **Congestive Heart Failure** |  |  |  |  | **0.05** |
| No | 117 (96.7%) | 151 (93.2%) | 164 (89.1%) | 432 (92.5%) |  |
| Yes | 4 (3.3%) | 11 (6.8%) | 20 (10.9%) | 35 (7.5%) |  |
| **Ejection Fraction** |  |  |  |  | 0.15 |
| Normal (>49%) | 110 (90.9%) | 147 (90.7%) | 162 (88.0%) | 419 (89.7%) |  |
| Reduced (<35%)/Moderately Reduced (35%-49%) | 2 (1.7%) | 4 (2.5%) | 10 (5.4%) | 16 (3.4%) |  |
| Missing | 9 (7.4%) | 11 (6.8%) | 12 (6.5%) | 32 (6.9%) |  |
| **Chronic Kidney Disease** |  |  |  |  | 0.3 |
| No | 79 (65.3%) | 116 (71.6%) | 118 (64.1%) | 313 (67.0%) |  |
| Yes | 42 (34.7%) | 46 (28.4%) | 66 (35.9%) | 154 (33.0%) |  |
| **Hyperlipidemia** |  |  |  |  | 0.2 |
| No | 19 (15.7%) | 37 (22.8%) | 44 (23.9%) | 100 (21.4%) |  |
| Yes | 102 (84.3%) | 125 (77.2%) | 140 (76.1%) | 367 (78.6%) |  |
| **Total Cholesterol** |  |  |  |  | 0.26 |
| Mean (SD) | 173 (43.0) | 178 (44.2) | 182 (43.5) | 178 (43.6) |  |
| Median [Min, Max] | 171 [86.0, 267] | 176 [46.0, 338] | 180 [94.0, 367] | 177 [46.0, 367] |  |
| Missing | 6 (5.0%) | 15 (9.3%) | 9 (4.9%) | 30 (6.4%) |  |
| **HDL** |  |  |  |  | 0.76 |
| Mean (SD) | 54.8 (22.3) | 52.9 (17.3) | 53.1 (27.2) | 53.5 (22.9) |  |
| Median [Min, Max] | 49.0 [26.0, 180] | 49.0 [23.0, 117] | 47.0 [20.0, 274] | 48.0 [20.0, 274] |  |
| Missing | 6 (5.0%) | 15 (9.3%) | 9 (4.9%) | 30 (6.4%) |  |
| **LDL** |  |  |  |  | 0.12 |
| Mean (SD) | 95.5 (36.5) | 104 (37.4) | 103 (36.8) | 102 (37.0) |  |
| Median [Min, Max] | 94.0 [25.0, 183] | 101 [35.0, 250] | 102 [29.0, 243] | 100 [25.0, 250] |  |
| Missing | 6 (5.0%) | 15 (9.3%) | 10 (5.4%) | 31 (6.6%) |  |
| **Triglycerides** |  |  |  |  | **0.02** |
| Mean (SD) | 115 (59.3) | 110 (56.7) | 130 (77.3) | 119 (66.8) |  |
| Median [Min, Max] | 102 [32.0, 336] | 100 [8.00, 346] | 112 [37.0, 435] | 102 [8.00, 435] |  |
| Missing | 6 (5.0%) | 17 (10.5%) | 12 (6.5%) | 35 (7.5%) |  |
| **Statin** |  |  |  |  | 0.9 |
| No | 44 (36.4%) | 63 (38.9%) | 72 (39.1%) | 179 (38.3%) |  |
| Yes | 77 (63.6%) | 99 (61.1%) | 112 (60.9%) | 288 (61.7%) |  |
| **Atrial Fibrillation** |  |  |  |  | 0.17 |
| No | 114 (94.2%) | 159 (98.1%) | 174 (94.6%) | 447 (95.7%) |  |
| Yes | 7 (5.8%) | 3 (1.9%) | 10 (5.4%) | 20 (4.3%) |  |
| **Anticoagulation** |  |  |  |  | 0.5 |
| No | 110 (90.9%) | 153 (94.4%) | 172 (93.5%) | 435 (93.1%) |  |
| Yes | 11 (9.1%) | 9 (5.6%) | 12 (6.5%) | 32 (6.9%) |  |
| **Coronary Artery Disease** |  |  |  |  | 0.41 |
| No | 107 (88.4%) | 142 (87.7%) | 154 (83.7%) | 403 (86.3%) |  |
| Yes | 14 (11.6%) | 20 (12.3%) | 30 (16.3%) | 64 (13.7%) |  |
| **Smoker** |  |  |  |  | 0.05 |
| No | 59 (48.8%) | 91 (56.2%) | 80 (43.5%) | 230 (49.3%) |  |
| Yes - Current | 22 (18.2%) | 33 (20.4%) | 53 (28.8%) | 108 (23.1%) |  |
| Yes - Former | 40 (33.1%) | 38 (23.5%) | 51 (27.7%) | 129 (27.6%) |  |
| **Prior Transient Ischemic Attack/Stroke/Subarachnoid Hemorrhage** |  |  |  |  | 0.97 |
| No | 115 (95.0%) | 154 (95.1%) | 174 (94.6%) | 443 (94.9%) |  |
| Yes | 6 (5.0%) | 8 (4.9%) | 10 (5.4%) | 24 (5.1%) |  |
| **Obesity (BMI > 30)** |  |  |  |  | 0.05 |
| No | 58 (47.9%) | 68 (42.0%) | 63 (34.2%) | 189 (40.5%) |  |
| Yes | 63 (52.1%) | 94 (58.0%) | 121 (65.8%) | 278 (59.5%) |  |
| **Aspirin Therapy** |  |  |  |  | 0.11 |
| No | 99 (81.8%) | 133 (82.1%) | 136 (73.9%) | 368 (78.8%) |  |
| Yes | 22 (18.2%) | 29 (17.9%) | 48 (26.1%) | 99 (21.2%) |  |
| <sup>1</sup> The ANOVA test is used for continuous variables, while the Chi-Square test is used for categorical variables. The p-value indicates whether the variable differs across the three blood pressure groups. |  |  |  |  |  |

Data are presented as mean (SD) for continuous variables and n (%) for categorical variables, stratified by blood pressure (BP) control status at the 3-month follow-up visit (Controlled, Moderately Controlled, Uncontrolled). BMI indicates body mass index; DBP, diastolic blood pressure; HDL, high-density lipoprotein; LDL, low-density lipoprotein; SBP, systolic blood pressure.

**Table S2b:** Summary of 6-Month Visit Data.

|  | **Controlled** | **Moderately Controlled** | **Uncontrolled** | **Overall** | **p-value <sup>1</sup>** |
| --- | --- | --- | --- | --- | --- |
|  | **(N=120)** | **(N=162)** | **(N=134)** | **(N=416)** |  |
| **Age** |  |  |  |  | **<0.001** |
| Mean (SD) | 62.0 (10.6) | 59.2 (12.4) | 56.3 (11.7) | 59.1 (11.9) |  |
| Median [Min, Max] | 63.0 [34.0, 87.0] | 59.0 [28.0, 86.0] | 57.0 [27.0, 77.0] | 60.0 [27.0, 87.0] |  |
| **Sex** |  |  |  |  | 0.91 |
| Female | 56 (46.7%) | 77 (47.5%) | 66 (49.3%) | 199 (47.8%) |  |
| Male | 64 (53.3%) | 85 (52.5%) | 68 (50.7%) | 217 (52.2%) |  |
| **Race** |  |  |  |  | **<0.001** |
| African-American | 51 (42.5%) | 105 (64.8%) | 109 (81.3%) | 265 (63.7%) |  |
| Other | 11 (9.2%) | 11 (6.8%) | 7 (5.2%) | 29 (7.0%) |  |
| White | 58 (48.3%) | 46 (28.4%) | 18 (13.4%) | 122 (29.3%) |  |
| **SBP** |  |  |  |  | **<0.001** |
| Mean (SD) | 118 (8.80) | 130 (5.92) | 149 (13.0) | 133 (15.7) |  |
| Median [Min, Max] | 121 [90.0, 129] | 131 [108, 139] | 147 [123, 196] | 131 [90.0, 196] |  |
| **DBP** |  |  |  |  | **<0.001** |
| Mean (SD) | 71.9 (6.04) | 81.3 (5.98) | 89.9 (11.5) | 81.3 (10.8) |  |
| Median [Min, Max] | 74.0 [50.0, 79.0] | 82.0 [54.0, 89.0] | 90.0 [54.0, 128] | 81.0 [50.0, 128] |  |
| **Number of Anti-Hypertensive Medications** |  |  |  |  | **<0.001** |
| 0 | 11 (9.2%) | 12 (7.4%) | 3 (2.2%) | 26 (6.3%) |  |
| 1 | 40 (33.3%) | 49 (30.2%) | 23 (17.2%) | 112 (26.9%) |  |
| 2 | 46 (38.3%) | 51 (31.5%) | 42 (31.3%) | 139 (33.4%) |  |
| 3 or more than 3 | 23 (19.2%) | 50 (30.9%) | 66 (49.3%) | 139 (33.4%) |  |
| **Type 2 Diabetes Mellitus** |  |  |  |  | 0.88 |
| No | 81 (67.5%) | 112 (69.1%) | 89 (66.4%) | 282 (67.8%) |  |
| Yes | 39 (32.5%) | 50 (30.9%) | 45 (33.6%) | 134 (32.2%) |  |
| **Congestive Heart Failure** |  |  |  |  | **0.01** |
| No | 115 (95.8%) | 154 (95.1%) | 117 (87.3%) | 386 (92.8%) |  |
| Yes | 5 (4.2%) | 8 (4.9%) | 17 (12.7%) | 30 (7.2%) |  |
| **Ejection Fraction** |  |  |  |  | 0.26 |
| Normal (>49%) | 110 (91.7%) | 146 (90.1%) | 122 (91.0%) | 378 (90.9%) |  |
| Reduced (<35%)/Moderately Reduced (35%-49%) | 3 (2.5%) | 3 (1.9%) | 7 (5.2%) | 13 (3.1%) |  |
| Missing | 7 (5.8%) | 13 (8.0%) | 5 (3.7%) | 25 (6.0%) |  |
| **Chronic Kidney Disease** |  |  |  |  | 0.36 |
| No | 83 (69.2%) | 109 (67.3%) | 82 (61.2%) | 274 (65.9%) |  |
| Yes | 37 (30.8%) | 53 (32.7%) | 52 (38.8%) | 142 (34.1%) |  |
| **Hyperlipidemia** |  |  |  |  | 0.96 |
| No | 25 (20.8%) | 32 (19.8%) | 26 (19.4%) | 83 (20.0%) |  |
| Yes | 95 (79.2%) | 130 (80.2%) | 108 (80.6%) | 333 (80.0%) |  |
| **Total Cholesterol** |  |  |  |  | 0.45 |
| Mean (SD) | 175 (44.6) | 182 (37.7) | 178 (48.4) | 178 (43.4) |  |
| Median [Min, Max] | 176 [86.0, 275] | 181 [73.0, 284] | 172 [46.0, 367] | 176 [46.0, 367] |  |
| Missing | 8 (6.7%) | 12 (7.4%) | 5 (3.7%) | 25 (6.0%) |  |
| **HDL** |  |  |  |  | 0.82 |
| Mean (SD) | 53.5 (21.7) | 54.5 (18.2) | 52.8 (28.4) | 53.6 (22.9) |  |
| Median [Min, Max] | 48.5 [23.0, 180] | 49.5 [25.0, 115] | 47.0 [20.0, 274] | 48.0 [20.0, 274] |  |
| Missing | 8 (6.7%) | 12 (7.4%) | 5 (3.7%) | 25 (6.0%) |  |
| **LDL** |  |  |  |  | 0.41 |
| Mean (SD) | 97.4 (37.9) | 103 (34.6) | 103 (39.6) | 101 (37.3) |  |
| Median [Min, Max] | 93.0 [25.0, 187] | 104 [35.0, 212] | 98.0 [29.0, 250] | 100 [25.0, 250] |  |
| Missing | 8 (6.7%) | 13 (8.0%) | 5 (3.7%) | 26 (6.3%) |  |
| **Triglycerides** |  |  |  |  | 0.58 |
| Mean (SD) | 120 (64.1) | 117 (61.3) | 125 (77.3) | 121 (67.7) |  |
| Median [Min, Max] | 102 [8.00, 394] | 106 [32.0, 336] | 104 [20.0, 407] | 103 [8.00, 407] |  |
| Missing | 8 (6.7%) | 14 (8.6%) | 7 (5.2%) | 29 (7.0%) |  |
| **Statin** |  |  |  |  | 0.63 |
| No | 41 (34.2%) | 63 (38.9%) | 46 (34.3%) | 150 (36.1%) |  |
| Yes | 79 (65.8%) | 99 (61.1%) | 88 (65.7%) | 266 (63.9%) |  |
| **Atrial Fibrillation** |  |  |  |  | 0.32 |
| No | 115 (95.8%) | 158 (97.5%) | 126 (94.0%) | 399 (95.9%) |  |
| Yes | 5 (4.2%) | 4 (2.5%) | 8 (6.0%) | 17 (4.1%) |  |
| **Anticoagulation** |  |  |  |  | 0.03 |
| No | 111 (92.5%) | 158 (97.5%) | 121 (90.3%) | 390 (93.8%) |  |
| Yes | 9 (7.5%) | 4 (2.5%) | 13 (9.7%) | 26 (6.3%) |  |
| **Coronary Artery Disease** |  |  |  |  | 0.1 |
| No | 107 (89.2%) | 142 (87.7%) | 108 (80.6%) | 357 (85.8%) |  |
| Yes | 13 (10.8%) | 20 (12.3%) | 26 (19.4%) | 59 (14.2%) |  |
| **Smoker** |  |  |  |  | 0.07 |
| No | 66 (55.0%) | 85 (52.5%) | 54 (40.3%) | 205 (49.3%) |  |
| Yes - Current | 20 (16.7%) | 35 (21.6%) | 40 (29.9%) | 95 (22.8%) |  |
| Yes - Former | 34 (28.3%) | 42 (25.9%) | 40 (29.9%) | 116 (27.9%) |  |
| **Prior Transient Ischemic Attack/Stroke/Subarachnoid Hemorrhage** |  |  |  |  | 0.76 |
| No | 115 (95.8%) | 152 (93.8%) | 127 (94.8%) | 394 (94.7%) |  |
| Yes | 5 (4.2%) | 10 (6.2%) | 7 (5.2%) | 22 (5.3%) |  |
| **Obesity (BMI > 30)** |  |  |  |  | 0.29 |
| No | 57 (47.5%) | 72 (44.4%) | 51 (38.1%) | 180 (43.3%) |  |
| Yes | 63 (52.5%) | 90 (55.6%) | 83 (61.9%) | 236 (56.7%) |  |
| **Aspirin Therapy** |  |  |  |  | 0.57 |
| No | 97 (80.8%) | 127 (78.4%) | 101 (75.4%) | 325 (78.1%) |  |
| Yes | 23 (19.2%) | 35 (21.6%) | 33 (24.6%) | 91 (21.9%) |  |
| <sup>1</sup> The ANOVA test is used for continuous variables, while the Chi-Square test is used for categorical variables. The p-value indicates whether the variable differs across the three blood pressure groups. |  |  |  |  |  |

Data are presented as mean (SD) for continuous variables and n (%) for categorical variables, stratified by blood pressure (BP) control status at the 6-month follow-up visit (Controlled, Moderately Controlled, Uncontrolled). BMI indicates body mass index; DBP, diastolic blood pressure; HDL, high-density lipoprotein; LDL, low-density lipoprotein; SBP, systolic blood pressure.

**Table S2c:** Summary of 9-Month Visit Data.

|  | **Controlled** | **Moderately Controlled** | **Uncontrolled** | **Overall** | **p-value <sup>1</sup>** |
| --- | --- | --- | --- | --- | --- |
|  | **(N=135)** | **(N=116)** | **(N=113)** | **(N=364)** |  |
| **Age** |  |  |  |  | **<0.001** |
| Mean (SD) | 62.1 (12.2) | 59.6 (10.4) | 56.3 (12.0) | 59.5 (11.8) |  |
| Median [Min, Max] | 64.0 [27.0, 86.0] | 59.5 [33.0, 87.0] | 58.0 [28.0, 83.0] | 60.5 [27.0, 87.0] |  |
| **Sex** |  |  |  |  | 0.21 |
| Female | 73 (54.1%) | 51 (44.0%) | 51 (45.1%) | 175 (48.1%) |  |
| Male | 62 (45.9%) | 65 (56.0%) | 62 (54.9%) | 189 (51.9%) |  |
| **Race** |  |  |  |  | **<0.001** |
| African-American | 70 (51.9%) | 78 (67.2%) | 89 (78.8%) | 237 (65.1%) |  |
| Other | 10 (7.4%) | 7 (6.0%) | 8 (7.1%) | 25 (6.9%) |  |
| White | 55 (40.7%) | 31 (26.7%) | 16 (14.2%) | 102 (28.0%) |  |
| **SBP** |  |  |  |  | **<0.001** |
| Mean (SD) | 119 (8.09) | 130 (6.47) | 150 (14.6) | 132 (16.3) |  |
| Median [Min, Max] | 121 [91.0, 129] | 131 [102, 139] | 147 [123, 224] | 129 [91.0, 224] |  |
| **DBP** |  |  |  |  | **<0.001** |
| Mean (SD) | 71.8 (5.75) | 80.7 (5.48) | 91.7 (13.9) | 80.8 (12.2) |  |
| Median [Min, Max] | 73.0 [46.0, 80.0] | 82.0 [59.0, 89.0] | 92.0 [63.0, 189] | 80.0 [46.0, 189] |  |
| **Number of Anti-Hypertensive Medications** |  |  |  |  | 0.03 |
| 0 | 14 (10.4%) | 9 (7.8%) | 5 (4.4%) | 28 (7.7%) |  |
| 1 | 39 (28.9%) | 40 (34.5%) | 22 (19.5%) | 101 (27.7%) |  |
| 2 | 36 (26.7%) | 36 (31.0%) | 35 (31.0%) | 107 (29.4%) |  |
| 3 or more than 3 | 46 (34.1%) | 31 (26.7%) | 51 (45.1%) | 128 (35.2%) |  |
| **Type 2 Diabetes Mellitus** |  |  |  |  | 0.48 |
| No | 86 (63.7%) | 79 (68.1%) | 80 (70.8%) | 245 (67.3%) |  |
| Yes | 49 (36.3%) | 37 (31.9%) | 33 (29.2%) | 119 (32.7%) |  |
| **Congestive Heart Failure** |  |  |  |  | 0.36 |
| No | 127 (94.1%) | 108 (93.1%) | 101 (89.4%) | 336 (92.3%) |  |
| Yes | 8 (5.9%) | 8 (6.9%) | 12 (10.6%) | 28 (7.7%) |  |
| **Ejection Fraction** |  |  |  |  | 0.7 |
| Normal (>49%) | 121 (89.6%) | 107 (92.2%) | 103 (91.2%) | 331 (90.9%) |  |
| Reduced (<35%)/Moderately Reduced (35%-49%) | 5 (3.7%) | 2 (1.7%) | 4 (3.5%) | 11 (3.0%) |  |
| Missing | 9 (6.7%) | 7 (6.0%) | 6 (5.3%) | 22 (6.0%) |  |
| **Chronic Kidney Disease** |  |  |  |  | 0.57 |
| No | 82 (60.7%) | 78 (67.2%) | 72 (63.7%) | 232 (63.7%) |  |
| Yes | 53 (39.3%) | 38 (32.8%) | 41 (36.3%) | 132 (36.3%) |  |
| **Hyperlipidemia** |  |  |  |  | 0.54 |
| No | 22 (16.3%) | 25 (21.6%) | 23 (20.4%) | 70 (19.2%) |  |
| Yes | 113 (83.7%) | 91 (78.4%) | 90 (79.6%) | 294 (80.8%) |  |
| **Total Cholesterol** |  |  |  |  | 0.12 |
| Mean (SD) | 174 (43.1) | 182 (40.6) | 186 (51.6) | 180 (45.3) |  |
| Median [Min, Max] | 176 [86.0, 268] | 180 [73.0, 305] | 181 [46.0, 367] | 180 [46.0, 367] |  |
| Missing | 6 (4.4%) | 8 (6.9%) | 8 (7.1%) | 22 (6.0%) |  |
| **HDL** |  |  |  |  | 0.47 |
| Mean (SD) | 54.0 (19.8) | 56.5 (20.8) | 52.6 (30.8) | 54.4 (24.0) |  |
| Median [Min, Max] | 50.0 [26.0, 180] | 51.0 [26.0, 147] | 46.0 [20.0, 274] | 49.0 [20.0, 274] |  |
| Missing | 6 (4.4%) | 8 (6.9%) | 8 (7.1%) | 22 (6.0%) |  |
| **LDL** |  |  |  |  | <0.01 |
| Mean (SD) | 96.6 (36.9) | 101 (36.9) | 113 (40.9) | 103 (38.6) |  |
| Median [Min, Max] | 93.0 [25.0, 189] | 96.5 [29.0, 212] | 108 [29.0, 250] | 101 [25.0, 250] |  |
| Missing | 6 (4.4%) | 8 (6.9%) | 9 (8.0%) | 23 (6.3%) |  |
| **Triglycerides** |  |  |  |  | 0.94 |
| Mean (SD) | 118 (59.5) | 119 (67.7) | 121 (67.8) | 119 (64.6) |  |
| Median [Min, Max] | 101 [20.0, 336] | 105 [8.00, 346] | 102 [36.0, 371] | 102 [8.00, 371] |  |
| Missing | 7 (5.2%) | 8 (6.9%) | 10 (8.8%) | 25 (6.9%) |  |
| **Statin** |  |  |  |  | 0.54 |
| No | 45 (33.3%) | 40 (34.5%) | 45 (39.8%) | 130 (35.7%) |  |
| Yes | 90 (66.7%) | 76 (65.5%) | 68 (60.2%) | 234 (64.3%) |  |
| **Atrial Fibrillation** |  |  |  |  | 0.95 |
| No | 130 (96.3%) | 111 (95.7%) | 108 (95.6%) | 349 (95.9%) |  |
| Yes | 5 (3.7%) | 5 (4.3%) | 5 (4.4%) | 15 (4.1%) |  |
| **Anticoagulation** |  |  |  |  | 0.16 |
| No | 126 (93.3%) | 112 (96.6%) | 102 (90.3%) | 340 (93.4%) |  |
| Yes | 9 (6.7%) | 4 (3.4%) | 11 (9.7%) | 24 (6.6%) |  |
| **Coronary Artery Disease** |  |  |  |  | 0.81 |
| No | 116 (85.9%) | 101 (87.1%) | 95 (84.1%) | 312 (85.7%) |  |
| Yes | 19 (14.1%) | 15 (12.9%) | 18 (15.9%) | 52 (14.3%) |  |
| **Smoker** |  |  |  |  | 0.4 |
| No | 63 (46.7%) | 58 (50.0%) | 51 (45.1%) | 172 (47.3%) |  |
| Yes - Current | 28 (20.7%) | 27 (23.3%) | 34 (30.1%) | 89 (24.5%) |  |
| Yes - Former | 44 (32.6%) | 31 (26.7%) | 28 (24.8%) | 103 (28.3%) |  |
| **Prior Transient Ischemic Attack/Stroke/Subarachnoid Hemorrhage** |  |  |  |  | 0.42 |
| No | 129 (95.6%) | 112 (96.6%) | 105 (92.9%) | 346 (95.1%) |  |
| Yes | 6 (4.4%) | 4 (3.4%) | 8 (7.1%) | 18 (4.9%) |  |
| **Obesity (BMI > 30)** |  |  |  |  | 0.14 |
| No | 66 (48.9%) | 50 (43.1%) | 41 (36.3%) | 157 (43.1%) |  |
| Yes | 69 (51.1%) | 66 (56.9%) | 72 (63.7%) | 207 (56.9%) |  |
| **Aspirin Therapy** |  |  |  |  | 0.79 |
| No | 107 (79.3%) | 92 (79.3%) | 86 (76.1%) | 285 (78.3%) |  |
| Yes | 28 (20.7%) | 24 (20.7%) | 27 (23.9%) | 79 (21.7%) |  |
| <sup>1</sup> The ANOVA test is used for continuous variables, while the Chi-Square test is used for categorical variables. The p-value indicates whether the variable differs across the three blood pressure groups. |  |  |  |  |  |

Data are presented as mean (SD) for continuous variables and n (%) for categorical variables, stratified by blood pressure (BP) control status at the 9-month follow-up visit (Controlled, Moderately Controlled, Uncontrolled). BMI indicates body mass index; DBP, diastolic blood pressure; HDL, high-density lipoprotein; LDL, low-density lipoprotein; SBP, systolic blood pressure.

**Table S3:**
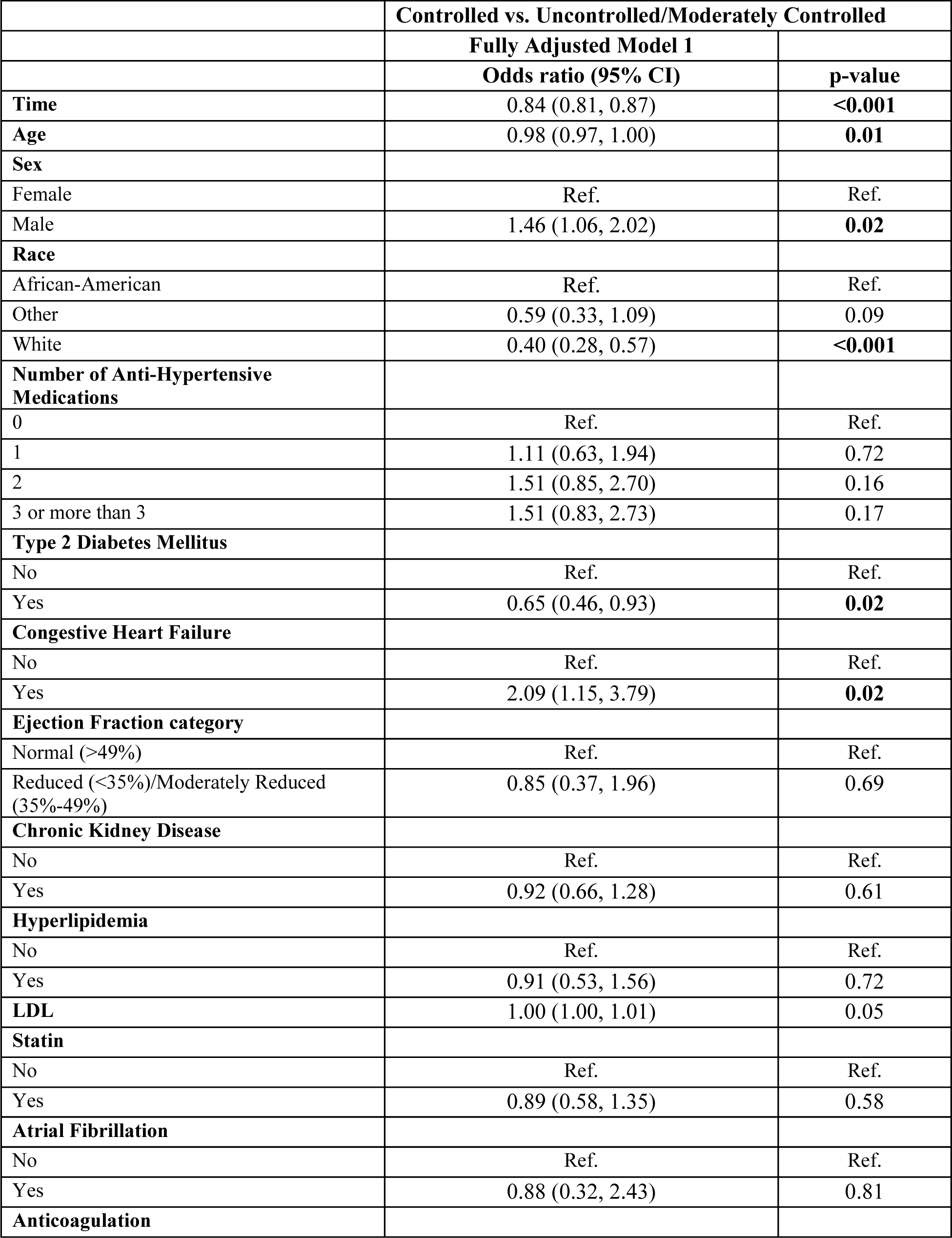
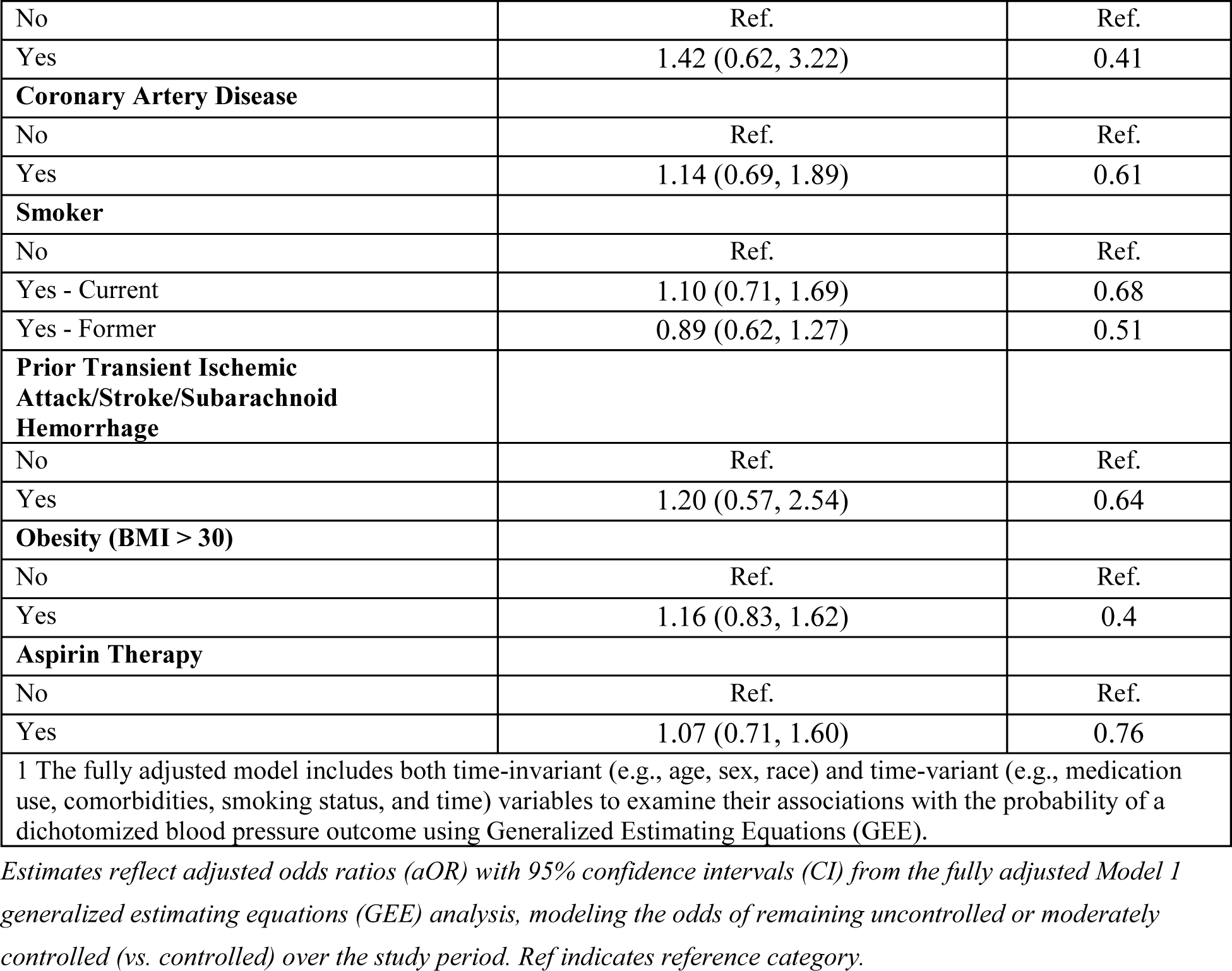
Longitudinal Analysis for Dichotomized Outcome (Controlled vs. Uncontrolled/Moderately Controlled, Ref:Controlled)

**Table S4:**
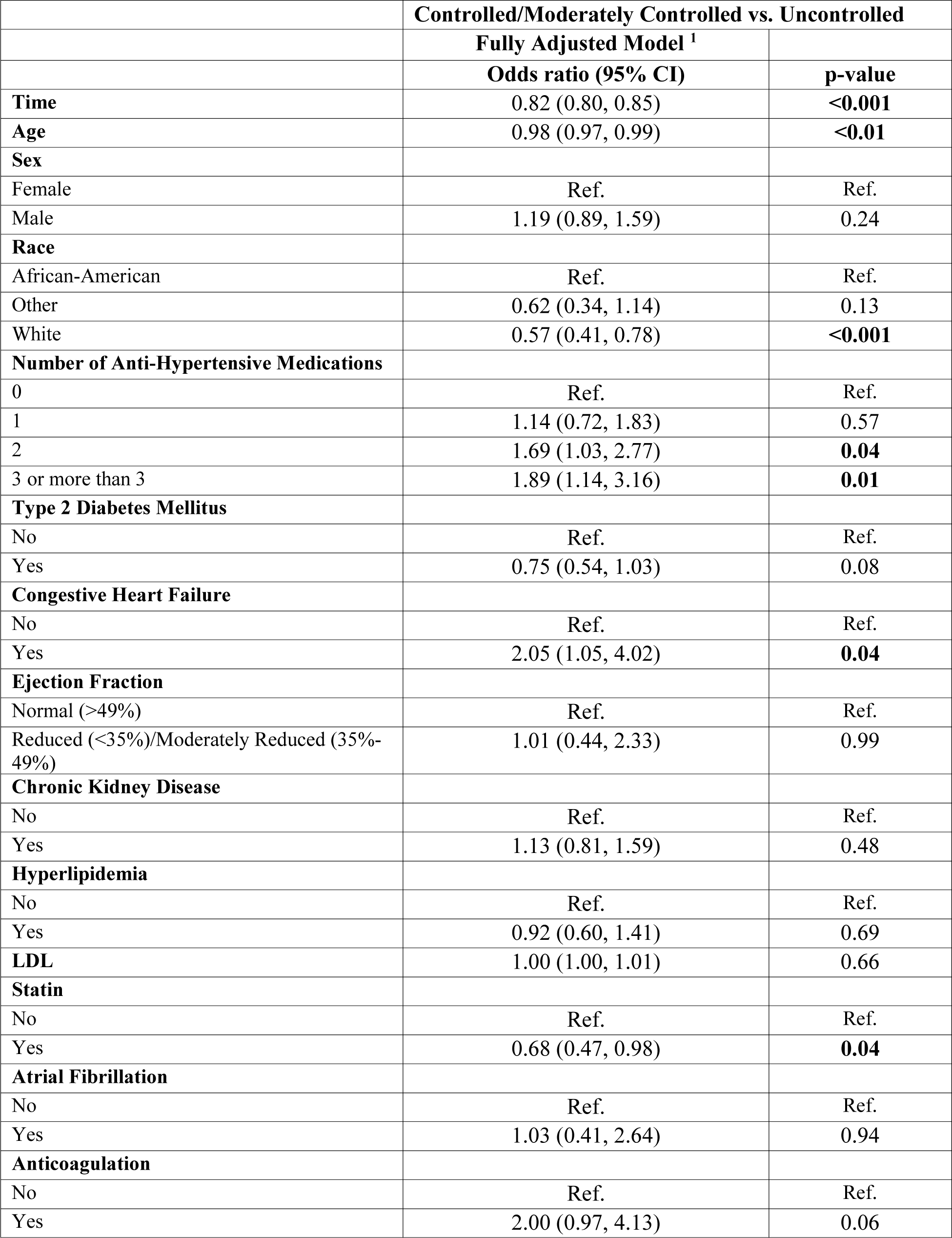
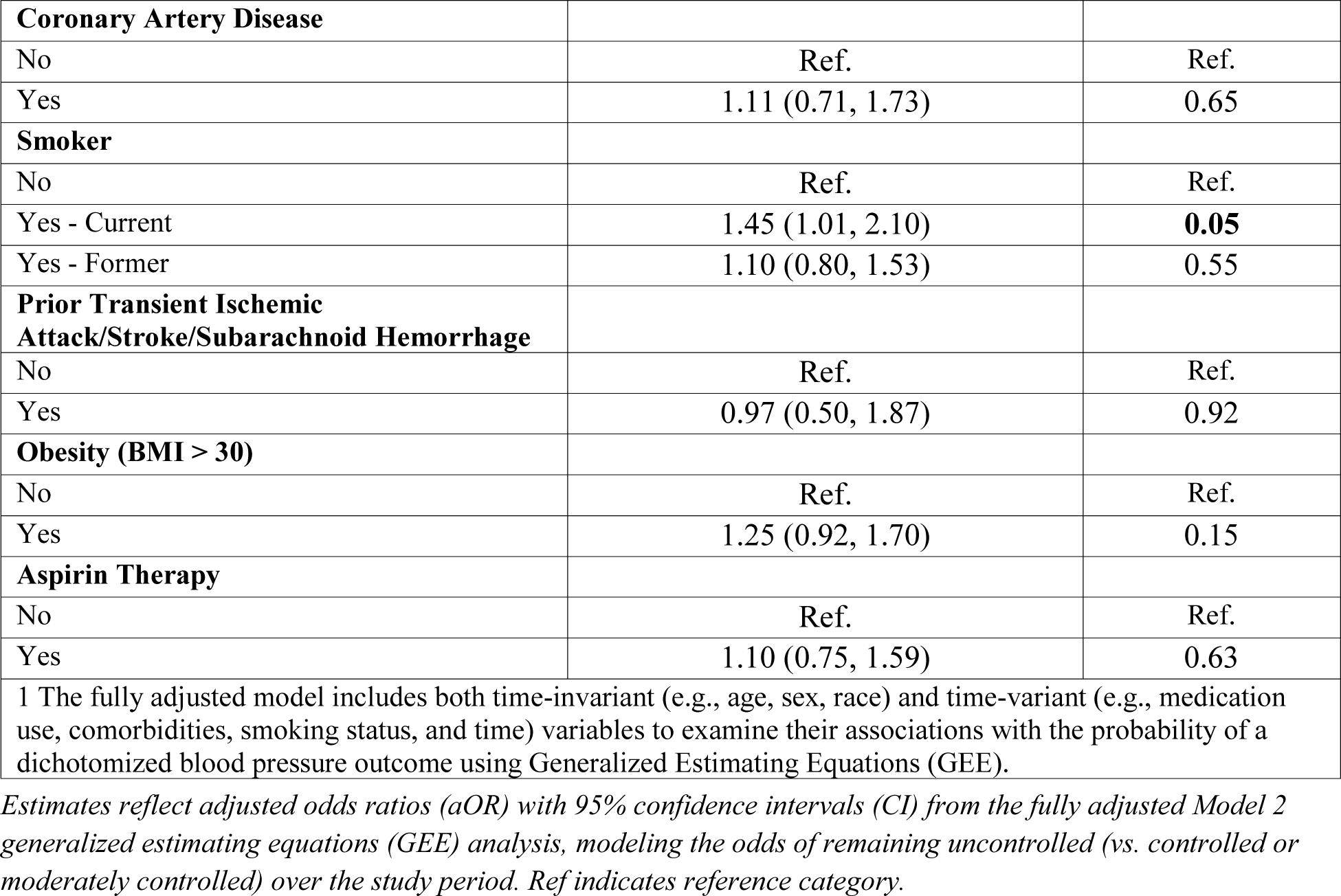
Longitudinal Analysis for Dichotomized Outcome (Controlled/Moderately Controlled vs. Uncontrolled, Ref:Controlled/Moderately Controlled)

## References

1. Whelton PK, Carey RM, Aronow WS, et al. 2017 ACC/AHA/AAPA/ABC/ACPM/AGS/APhA/ASH/ASPC/NMA/PCNA Guideline for the Prevention, Detection, Evaluation, and Management of High Blood Pressure in Adults. Circulation. 2018;138(17):e484–e594. doi:10.1161/CIR.0000000000000596

2. Mills KT, Stefanescu A, He J. The global epidemiology of hypertension. Nat Rev Nephrol. 2020;16(4):223–237. doi:10.1038/s41581-019-0244-2

3. Okonofua EC, Simpson KN, Jesri A, Rehman SU, Durkalski VL, Egan BM. Therapeutic inertia is an impediment to achieving the Healthy People 2010 blood pressure control goals. Hypertension. 2006;47(3):345–351. doi:10.1161/01.HYP.0000200702.76436.4b

4. Himmelfarb CRD, Commodore-Mensah Y, Hill MN. Expanding the role of nurses to improve hypertension care and control globally. Ann Glob Health. 2016;82(2):243–253. doi:10.1016/j.aogh.2016.02.004

5. Ferdinand KC, Yadav K, Nasser SA, et al. Disparities in hypertension and cardiovascular disease in Blacks: the critical role of medication adherence. J Clin Hypertens (Greenwich). 2017;19(10):1015–1024. doi:10.1111/jch.13089

6. Alvidrez J, Castille D, Laude-Sharp M, Rosario A, Tabor D. The National Institute on Minority Health and Health Disparities Research Framework. Am J Public Health. 2019;109(S1):S16–S20. doi:10.2105/AJPH.2018.304883

7. Margolis KL, Asche SE, Bergdall AR, et al. Effect of home blood pressure telemonitoring and pharmacist management on blood pressure control: a cluster randomized clinical trial. JAMA. 2013;310(1):46–56. doi:10.1001/jama.2013.6549

8. Mehta SJ, Volpp KG, Troxel AB, et al. Remote blood pressure monitoring with social support for patients with hypertension: a randomized clinical trial. JAMA Netw Open. 2024;7(6):e2413515. doi:10.1001/jamanetworkopen.2024.13515

9. Magnussen C, Ojeda FM, Leong DP, et al. Global effect of modifiable risk factors on cardiovascular disease and mortality. N Engl J Med. 2023;389(14):1273–1285. doi:10.1056/NEJMoa2206916

10. von Elm E, Altman DG, Egger M, et al. The Strengthening the Reporting of Observational Studies in Epidemiology (STROBE) statement: guidelines for reporting observational studies. Ann Intern Med. 2007;147(8):573–577. doi:10.7326/0003-4819-147-8-200710160-00010

11. Omboni S, McManus RJ, Bosworth HB, et al. Evidence and recommendations on the use of telemedicine for the management of arterial hypertension: an international expert position paper. Hypertension. 2020;76(5):1368–1383. doi:10.1161/HYPERTENSIONAHA.120.15873

12. US Department of Health and Human Services Office of Minority Health. National Hypertension Control Initiative: Addressing Disparities among Racial and Ethnic Minority Populations. Published June 2021. https://minorityhealth.hhs.gov/national-hypertension-control-initiative

13. Krasner AH, Hwang SJ, Levy D, Hamburg NM, Mitchell GF. Sex-related differences in drug treatment, prevalence and blood pressure control in primary care. J Hypertens. 2023;41(8):1265–1273. doi:10.1097/HJH.0000000000003455

14. Kim BJ, Lee H, Ku J, et al. Gender difference of blood pressure control rate and clinical prognosis in patients with resistant hypertension: real-world observation study. J Am Heart Assoc. 2023;12(9):e027619. doi:10.1161/JAHA.122.027619

15. Gillis EE, Sullivan JC. Sex differences in the renin-angiotensin-aldosterone system and its roles in hypertension, cardiovascular, and kidney diseases. Pharmaceuticals (Basel). 2023;16(5):689. doi:10.3390/ph16050689

16. Messerli FH, Hofstetter L, Agabiti-Rosei E, et al. Expertise: blood pressure and heart failure. Clin Hypertens (Greenwich). 2019;21(S1):S15–S23. doi:10.1186/s40885-019-0132-x

17. Gheorghiade M, Follath F, Ponikowski P, et al. Assessing and grading congestion in acute heart failure: a scientific statement from the acute heart failure committee of the Heart Failure Association of the European Society of Cardiology and endorsed by the European Society of Intensive Care Medicine. Eur J Heart Fail. 2010;12(5):423–433. doi:10.1093/eurjhf/hfq045

18. Shchekochikhin D, Al Ammary F, Lindenfeld J, Schrier R. Role of diuretics and ultrafiltration in congestive heart failure. Pharmaceuticals (Basel). 2013;6(7):851–866. doi:10.3390/ph6070851

19. Yancy CW, Jessup M, Bozkurt B, et al. 2022 AHA/ACC/HFSA Guideline for the Management of Heart Failure. J Am Coll Cardiol. 2022;79(17):e263–e421. doi:10.1016/j.jacc.2021.12.012

